# Early identification of rare disease using deep phenotyping of electronic health records

**DOI:** 10.64898/2026.09.11.26362811

**Authors:** Tate Tunstall, Pauline C. Ng, Lindsay Meyers, Maria-Renee Coldagelli, Lu Yang, Naveen Pereira, Mayowa Osundiji, Kate Im, Akash Kumar, Matthew Rabinowitz

**Author notes:** contributed equally to the work (Tate Tunstall, Pauline C. Ng). jointly supervised the work (Akash Kumar, Matthew Rabinowitz).

## Abstract

Patients with rare genetic disorders often wait years for a correct diagnosis, highlighting the urgent need for early and efficient triage for those at risk. Electronic health records (EHRs) are rich in useful information, yet the extent to which this data is sufficient to generate prediagnostic signals is uncertain. We trained machine learning models on billing codes and phenotypes extracted from clinical notes and applied these across eight conditions in a longitudinal cohort of roughly 3 million patient records from the Mayo Clinic, seeking early identification of conditions. The number of patients identified early varied by condition, ranging from 10% to 89% at 99% specificity. Median lead times exceeded one year prior to the first diagnostic billing code for most conditions, including Fabry disease (1,408 days), hereditary angioedema (2,755 days), neurofibromatosis type 1 (1,067 days), and hereditary hemorrhagic telangiectasia (2,320 days). We show that ML models trained on EHR data can prioritize patients for clinician review and confirmatory genetic testing and that the integration of diverse phenotypic data sources provides superior predictive value.

## Main

Rare diseases collectively affect an estimated 25-30 million Americans, yet individual conditions often remain undiagnosed for years. The diagnostic odyssey is a significant burden for patients and healthcare systems alike. In the United States, patients with rare diseases visit an average of eight physicians over more than seven years before receiving an accurate diagnosis ^1^, leading to delayed treatment, disease progression, and substantial psychological and financial costs ^2^.

Medical records contain myriad forms of largely untapped data. The systematic analysis of billing codes, clinical notes, laboratory results, and other structured and unstructured data offers an unprecedented opportunity to identify patients who may benefit from earlier diagnostic evaluation. Recent advances in large language models enable the extraction and integration of complex phenotypic information from these diverse data sources, especially from free text. Free text sources such as clinical notes capture a richer, more accurate picture of a patient’s health than ICD billing codes. Clinical notes document the nuanced, granular realities of patient care, including symptom severity, disease progression, and complex behavioral factors. By contrast, ICD codes are primarily administrative tools engineered for insurance reimbursement. This intrinsic mismatch means that highly specific phenotypes or adverse events often lack a dedicated billing code, forcing coders to use generic approximations that strip away critical clinical context. Furthermore, the administrative burden and delay associated with the billing process frequently result in documentation omissions, leaving structured billing data as an incomplete subset of the comprehensive clinical reality preserved in the unstructured text ^3^.

The construction of phenotype risk scores (PheRS) is one approach to quantifying an individual’s disease risk using information extracted from the EHR. By weighting observed clinical features according to their specificity for particular conditions, PheRS can identify individuals with phenotypic profiles suggestive of Mendelian disorders ^4^. However, these risk scores tend to increase after the initial suspicion of disease, a phenomenon known as “diagnostic convergence” ^5^, and therefore are limited in utility for catching disease significantly before clinical suspicion. Furthermore, PheRS are constructed using billing codes, and will miss additional information in other forms such as clinical notes.

Here, we leverage data from a cohort of over 3 million patients from the Mayo Clinic to evaluate how combining phenotype risk scores with additional phenotypic information extracted from clinical notes can identify a significant number of rare disease patients much earlier in their diagnostic odyssey. Our goal is to identify patients who merit earlier specialist evaluation or targeted genetic testing to facilitate diagnostic processes and not to replace clinical judgment We selected eight Mendelian rare diseases for analysis, spanning multiple organ systems and representing a diverse range of inheritance patterns, ages of onset, and clinical presentations. We focused on rare diseases that are detected in childhood or later, because these patients likely have long diagnostic odysseys and would have rich medical records.

- Hereditary hemorrhagic telangiectasia (HHT)
- Loeys-Dietz syndrome
- Neurofibromatosis type 1
- Hereditary angioedema types 1 and 2
- Noonan syndrome
- Fabry disease
- Charcot-Marie-Tooth disease
- Wilson disease

Together, these eight diseases span a spectrum of phenotypic distinctiveness, variable penetrance, and diagnostic complexity, allowing evaluation of model performance across heterogeneous real-world scenarios.

We identified rare disease patients using the billing codes (ICD-10) corresponding to each disease and used three criteria to determine potential diagnosis points: 1) date of the first occurrence of diagnostic ICD code; 2) date of clinical suspicion; and 3) date of first diagnostic genetic test. We extracted all available clinical notes and billing codes, and then applied a large language model (LLM) pipeline to extract Human Phenotype Ontology (HPO) terms from clinical notes, using both a well-established method, ClinPhen (^6^) and a custom-built pipeline using Gemini. A superset of detected HPO terms were then combined with phenotypic risk scores in our model. We also generated HPO terms and risk scores for age-matched control patients with no billing codes corresponding to rare disease (see Methods). After model training, we determined a risk threshold corresponding to 99% specificity in our control group. We then applied our model to all available timepoints in a well-curated test set of rare disease patients, using our 99% specificity threshold to determine disease detection.

For all diagnosis endpoints, we detected a significant portion of patients early: 43-100% prior to billing code diagnosis, 10-88% prior to the date of initial clinical suspicion, and 8-33% prior to the date of first genetic test(Table 1). Among patients detected early and across all potential diagnosis endpoints, lead time before diagnosis ranged from 279 to 5014 days. For rare disease patients, date of clinical suspicion is likely the most clinically relevant endpoint, given that non-specific genetic tests may be ordered early in the diagnostic odyssey and billing codes may appear well after clinical suspicion. Using the date of clinical suspicion as the endpoint, our model identified rare disease patients from 471 days (Wilson disease) to 2,755 days (hereditary angioedema) earlier (Fig. 2).

**Table 1.** Total patient count for each disease, and percentage of patients detected early at 99% specificity for the 3 different clinical endpoints. Before Diagnosis: date when diagnostic ICD code first appears in EHR, Before Mention: date when disease in first mentioned in clinical record, Before Genetic Testing: date when first genetic test was ordered

| Disease | Total Patient Count | Before Diagnosis | Before Mention | Before Genetic Testing |
| --- | --- | --- | --- | --- |
| Fabry Disease | 18 | 100 | 88.9 | 33.3 |
| Hereditary Angioedema Type 1 2 | 17 | 94.1 | 88.2 | 29.4 |
| Neurofibromatosis Type 1 | 65 | 87.7 | 75.4 | 15.4 |
| Loeys Dietz Syndrome | 10 | 85 | 60 | 10 |
| Noonan Syndrome | 15 | 80 | 60 | 10 |
| Hereditary Hemorrhagic Telangiectasia | 20 | 86.7 | 60 | 26.7 |
| Wilson Disease | 37 | 32.4 | 29.7 | 21.6 |
| Charcot-Marie-Tooth Disease | 58 | 43.1 | 10.3 | 8.6 |

**Fig 1:**
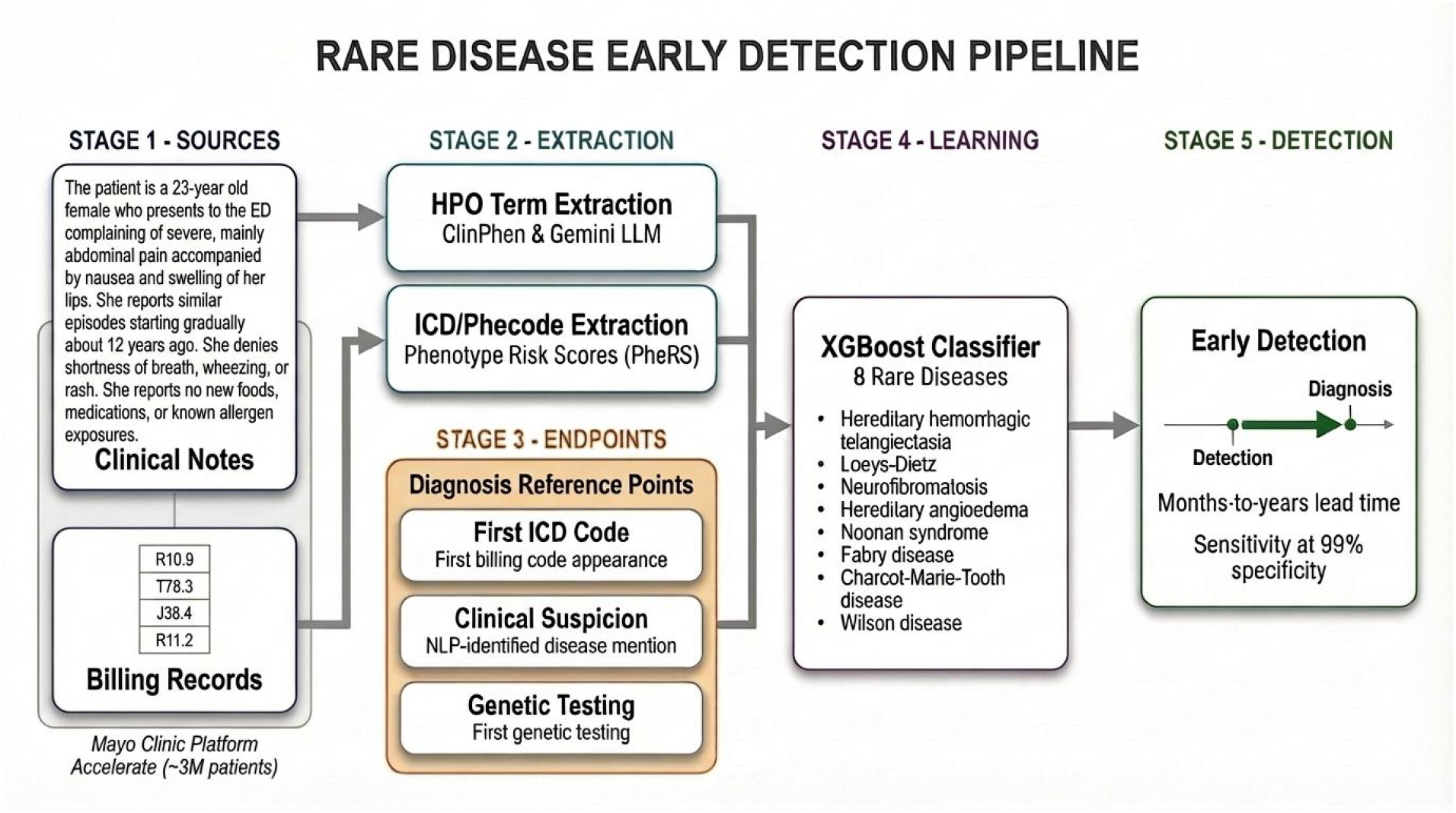
Rare disease early detection pipeline. HPO terms, ICD codes, and three potential diagnostic endpoints are extracted from electronic health records, which are then incorporated into a model predicting rare disease status at a threshold of 99% specificity

**Fig 2:**
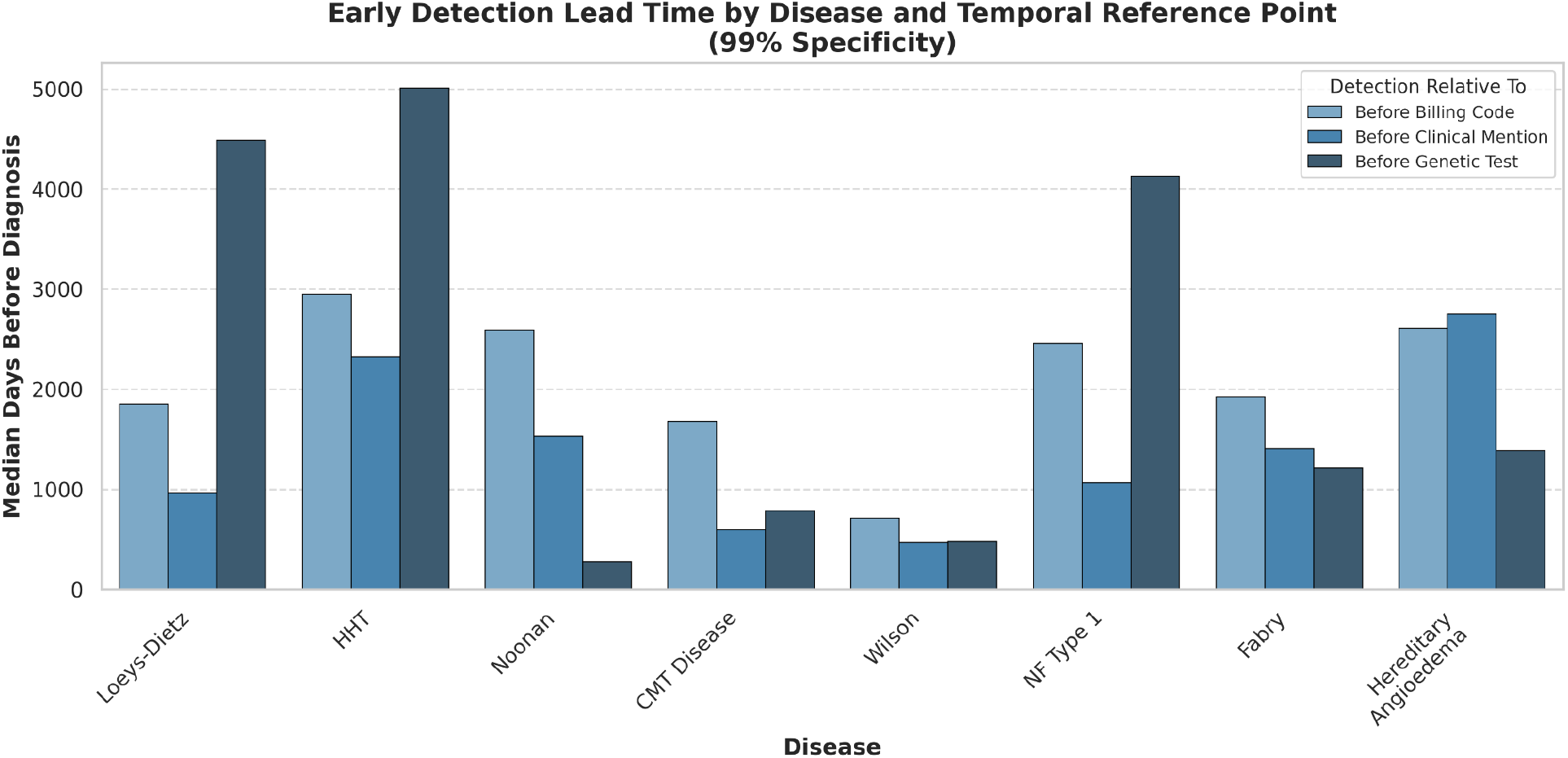
Median days before diagnosis before diagnosis for patients identified early. From left to right: Days before corresponding billing code first appears in patients EHR, days before disease was first mentioned in patients clinical notes, days before first record of genetic testing in patient’s EHR

Our success in early identification creates the opportunity for substantially accelerated diagnosis, intervention, and treatment. For many of the diseases studied, early detection can lead to significant improvement in patient care. For example, there are several long-term prophylactic therapies for hereditary angioedema such as treatment with monoclonal antibodies or RNA-targeted therapies ^7^. Early intervention and screening could lead to both the prevention and reduced progression of swelling attacks.

We assessed model interpretability using SHAP (SHapley Additive exPlanations) (Fig. 3). For all diseases except for neurofibromatosis and Fabry disease, feature importance analysis consistently identified the phenotype risk score as a top-ranking feature. However, the relative importance of the PheRS was moderate, indicating that HPO terms derived from clinical notes provided substantial additional discriminatory information compared to the billing code-derived risk scores. Our analysis consistently identified features clinically relevant to each disease, such as pneumothorax in Loeys–Dietz syndrome, neurofibromas in neurofibromatosis and foot weakness in Charcot-Marie-Tooth disease.

**Fig 3:**
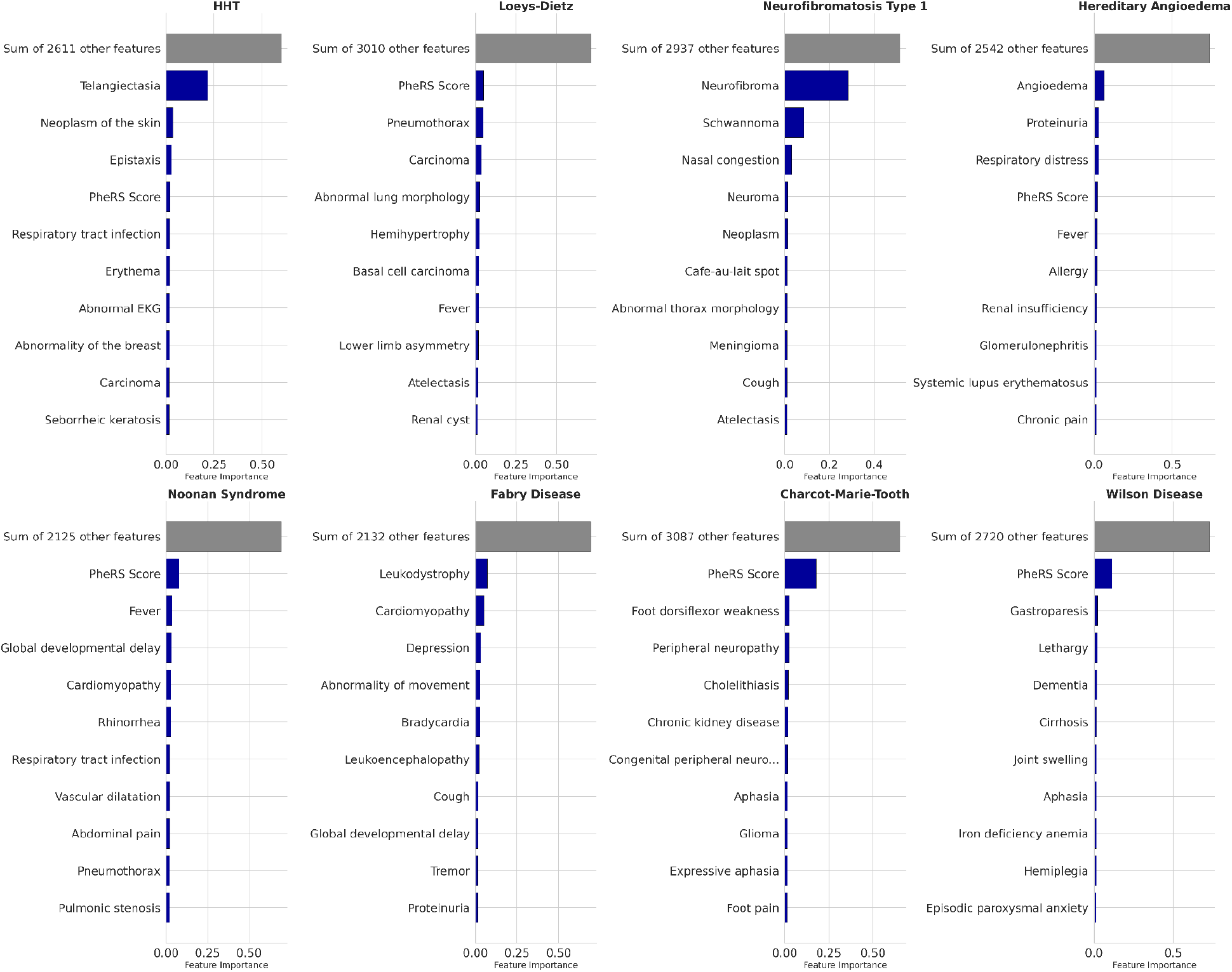
SHAP feature importance. Mean SHAP values for the top 10 most important features for each disease in blue. Grey bars indicate the total mean SHAP importance for the remaining features

The variation in early detection rates across diseases likely reflects differences in phenotypic expressivity, age of onset, and the distinctiveness of early disease manifestations. Fabry disease and hereditary angioedema, which showed the highest early detection rates using the clinical suspicion endpoint (89% and 88%, respectively), are characterized by relatively specific symptom constellations that may accumulate in the EHR prior to diagnostic consideration. In contrast, Charcot-Marie-Tooth disease, with the lowest early detection rate (10%), presents with more common and nonspecific neurological findings that overlap substantially with other conditions.

The viability of rule-based methods for identifying patients with rare disease at high specificity ^8^ is known from recent work. We show here that ML-based methods using a patient’s full EHR can identify patients years before a typical clinical diagnosis while maintaining a similar false-positive rate. Furthermore, while rule-based methods are inherently limited to a relatively small number of features, ML methods can integrate thousands of subtle features into a cohesive diagnostic signature. The consistent identification of clinically relevant features through feature importance analysis suggests that the models are learning genuine phenotypic relationships rather than spurious correlations. Furthermore, phenotype risk scores emerged as an important feature for most diseases, but never dominated the model, indicating that the integration of multiple, diverse phenotypic data sources provides value beyond any single approach.

Our work has several limitations. First, we studied a small subset of thousands of potential rare diseases. We selected diseases that typically present in adults, resulting in longer mean diagnosis times and more opportunity for early detection. Second, we cannot completely rule out the possibility that some patients are diagnosed within a different health system before transferring to the Mayo Clinic, resulting in the absence of corresponding diagnostic billing codes within the Mayo system. We tried to ameliorate this by carefully reviewing patient notes for mention of each rare disease. Finally, given the rarity of the diseases studied, even a specificity of 99% can yield relatively low precision, resulting in an excess of false-positive predictions. Individuals identified as at-risk for rare disease could follow up with low cost interventions such as visiting a specialist and collecting confirmatory phenotypic data, but the actual clinical and economic impact of system-wide monitoring for rare disease merits further study.

Our findings establish the foundation for EHR-based rare disease screening systems that could shorten the diagnostic odyssey experienced by millions of patients with undiagnosed rare conditions, and we believe this approach is scalable to many diverse health systems. An automated, low cost and high precision rare disease screening system could relieve pressure on primary care providers, prevent unnecessary treatments, and lead to significant cost savings by providing patients with preventative or ameliorative care. Future work integrating genomic data, advanced NLP methods, and real-time clinical decision support could translate these research findings into clinical practice, ultimately improving outcomes for patients with rare diseases.

## Methods

### Study Population

We used EHR data from Mayo Clinic, encompassing approximately 3 million patients with longitudinal clinical records including demographics, billing codes, clinical notes, laboratory results, and procedure records.

### Case Identification

Rare disease patients were identified based on the presence of ICD-10 billing codes corresponding to each target disease. For each condition, we selected the ICD-10 code or combination of codes that most accurately captured the diagnosis. Patients were required to have at least one occurrence of the relevant diagnostic code to be classified as cases.

### Control Selection

Control individuals were selected using stringent matching criteria to minimize confounding. Controls:

- Age-matched to cases (birthdate within ±1 year)
- Gender-matched to cases
- Had a clinical visit within ±2 months of the case’s first disease mention date
- Had no ICD codes corresponding to the rare diseases catalogued in the Mondo Disease Ontology rare disease list (https://mondo.monarchinitiative.org/pages/rare-disease/)

This strict matching protocol ensured that controls had comparable healthcare utilization patterns while excluding individuals with potential undiagnosed rare diseases.

### Diagnosis Endpoints

For each rare disease patient, we established the billing code diagnosis date as the earliest occurrence of the disease-specific ICD-10 code in the patient’s record. This date served as the reference point for evaluating early detection.

To establish a more clinically meaningful diagnosis date, we employed natural language processing to identify the first documented clinical suspicion of disease in the clinical notes. We searched for disease-specific terminology in conjunction with qualifying phrases indicating clinical suspicion, including: “possible,” “consistent,” “likely,” “screen,” “could,” “suspicious,” “investigative,” “could represent,” “diagnostic of,” “positive,” “probable,” “appearance of,” “seems to be,” and “impression.” The sentences containing these key words were inspected manually. If the sentence was part of a patient history, a referral, or family history, then this patient was discarded. We kept patients and first-suspicion dates where we were confident that this was the clinician’s first time suspecting the disease. For example, sentences like “This patient probably has Noonan syndrome” or “Ehlers-Danlos is possible.”

All NLP-identified dates of first clinical suspicion underwent manual chart review to confirm accuracy.

As a final more conservative endpoint, we recorded the earliest date of any relevant laboratory genetic test.

### Phenotype Risk Scores

Phenotype risk scores were generated using PheRS (Aref-Eshghi et al., 2022). The PheRS algorithm computes a weighted sum of an individual’s observed phecodes, where weights correspond to the log inverse prevalence of each phecode in the general population. This weighting scheme assigns higher scores to rare clinical features, reflecting the principle that unusual phenotypes are more informative for rare disease identification.

For each target disease, disease-specific PheRS were calculated based on the established phecode-to-disease mappings. The resulting scores quantify the degree of phenotypic overlap between an individual’s clinical presentation and the expected manifestations of each Mendelian condition.

### Feature Engineering

We extracted HPO terms from clinical notes using two methods:

1. **ClinPhen**: An automated clinical phenotype extraction tool that maps clinical text to standardized HPO terminology
2. **Gemini**: A large language model-based extraction system for identifying phenotypic features from unstructured clinical narratives

For each time point in a patient’s clinical record all HPO terms up until that point were one-hot-encoded into a representation of whether that phenotype had ever occurred in the patient’s record.

Phenotypic risk scores and HPO terms were then combined into a single vector representing all billing codes and phenotypes found in a patient’s record up to a given time point. For example if a patient suspected of having Fabry disease had previously experienced Obstructive hypertrophic cardiomyopathy (ICD10 code I42.1), this would be recorded as phecode 425.11 and given the corresponding weight in the pheRs package. This would then be combined with HPO terms found in clinical notes such as “abnormality of movement” (HP:0100022) or “leukodystrophy” (HP:0002415) as inputs to the model.

### Machine Learning Approach

Models were trained using gradient-boosted decision trees (XGBoost) with the number of boosting rounds set to 100 and a learning rate of 0.1. Models were trained on approximately 400 control patients, as described above, with no ICD codes corresponding to rare disease, and between 210-,1495 rare disease patients (see Sup Table 1 for per disease counts). Test data consisted of rare disease patients manually reviewed for rare disease status (see Diagnosis Endpoints and Table 1 for patient counts)

### Feature Importance

The Tree SHAP method ^9^ was used to compute exact SHAP values for each feature, providing both global and local explanations of feature contributions to model predictions.

## Data Availability

Data shown and reported in this manuscript have been extracted from this environment using an established protocol for data extraction, aimed at preserving patient privacy. The data have been de-identified pursuant to an expert determination in accordance with the HIPAA Privacy Rule. Any data beyond what is reported in the manuscript, including but not limited to the raw EHR data, cannot be shared or released due to the parameters of the expert determination to maintain the data de-identification.

## Acknowledgments

We would like to thank Adam Resnick and Nasibeh Zanjirani Farahani for their helpful advice on accessing the Mayo database. Stephen Montgomery provided helpful feedback on the manuscript.

## Author Contributions

T. Tunstall and P. Ng designed and carried out the study and prepared the manuscript. L. Meyers and M.R. Coldagelli contributed to disease selection and manuscript preparation. L. Yang assisted with phenotype extraction and pipeline design. N. Pereira and M. Osundidji contributed to rare disease phenotype selection and manuscript preparation, K. Im, A. Kumar and M. Rabinowitz contributed to study design and manuscript preparation.

## Data Availability

This study involves analysis of de-identified data via the Mayo Clinic Platform_Discover. In accordance with the Code of Federal Regulations, 45 CFR 46.102, the noted activity does not require IRB review. Data shown and reported in this manuscript have been extracted from this environment using an established protocol for data extraction, aimed at preserving patient privacy. The data have been de-identified pursuant to an expert determination in accordance with the HIPAA Privacy Rule. Any data beyond what is reported in the manuscript, including but not limited to the raw EHR data, cannot be shared or released due to the parameters of the expert determination to maintain the data de-identification.

**Supplemental table 1:** Training data patient count for each disease.

| Disease | Patient_Count | True_pos | True_neg |
| --- | --- | --- | --- |
| Fabry Disease | 801 | 389 | 412 |
| Hereditary Angioedema Type 1 2 | 1169 | 757 | 412 |
| Neurofibromatosis Type 1 | 1756 | 1344 | 412 |
| Loeys Dietz Syndrome | 1085 | 673 | 412 |
| Noonan Syndrome | 642 | 230 | 412 |
| Hereditary Hemorrhagic Telangiectasia | 1237 | 825 | 412 |
| Wilson Disease | 1150 | 738 | 412 |
| Charcot-Marie-Tooth Disease | 1907 | 1495 | 412 |

**Supplemental table 2:**
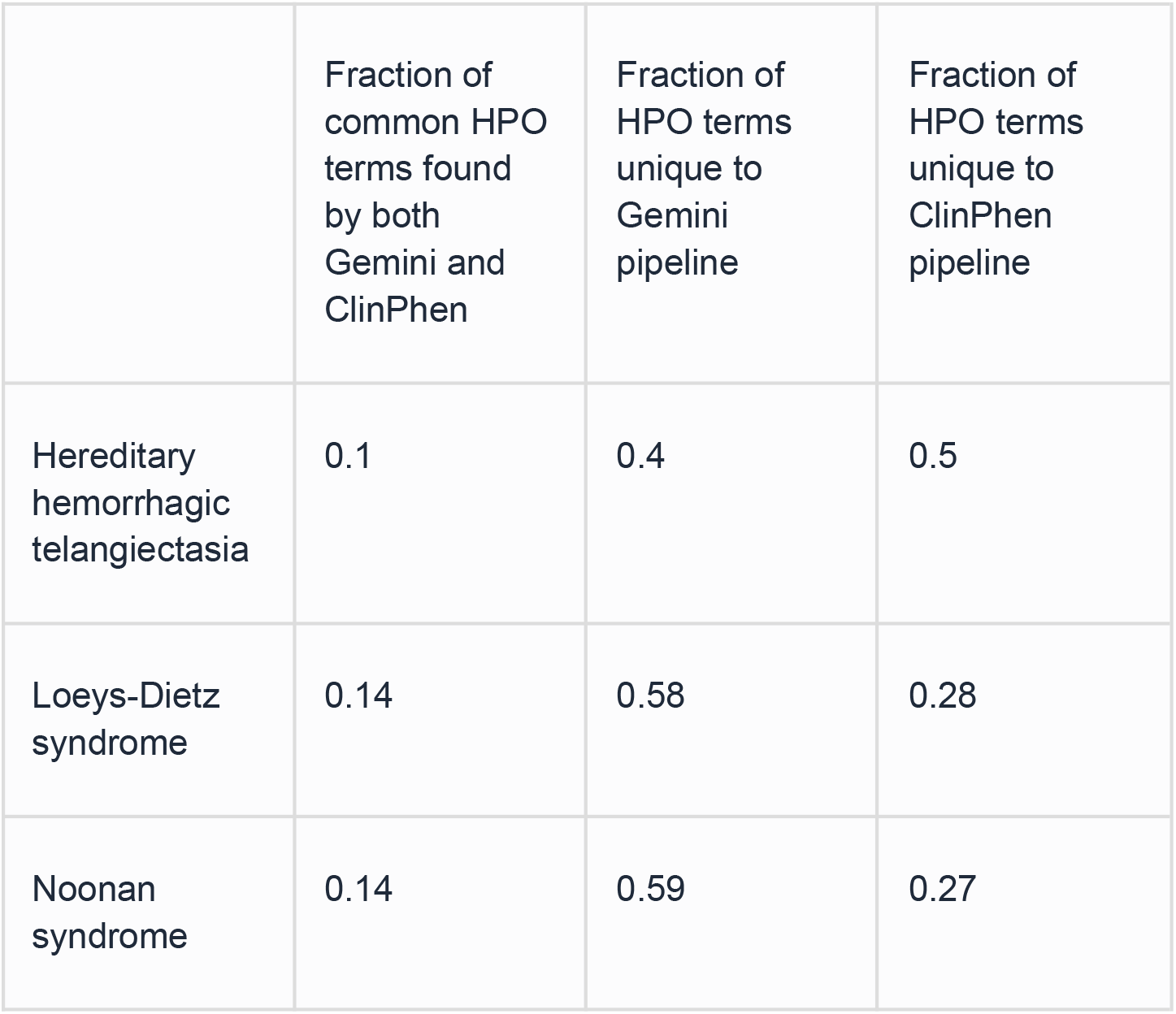
Comparison of LLM HPO extraction methods.

## References

1. Rare Disease Impact Report: Insights from Patients and the Medical Community. https://globalgenes.org/wp-content/uploads/2013/04/ShireReport-1.pdf (2013).

2. Tisdale, A. et al. The IDeaS initiative: pilot study to assess the impact of rare diseases on patients and healthcare systems. Orphanet J. Rare Dis. 16, 429 (2021).

3. Wei, W.-Q. et al. Combining billing codes, clinical notes, and medications from electronic health records provides superior phenotyping performance. J. Am. Med. Inform. Assoc. 23, e20–7 (2016).

4. Bastarache, L. et al. Phenotype risk scores identify patients with unrecognized Mendelian disease patterns. Science 359, 1233–1239 (2018).

5. Tinker, R. J., Peterson, J. & Bastarache, L. Phenotypic presentation of Mendelian disease across the diagnostic trajectory in electronic health records. Genet. Med. 25, 100921 (2023).

6. Deisseroth, C. A. et al. ClinPhen extracts and prioritizes patient phenotypes directly from medical records to expedite genetic disease diagnosis. Genet. Med. 21, 1585–1593 (2019).

7. Uminski, K., Goodyear, D. & Betschel, S. Therapeutic advances in hereditary angioedema: A focus on present and future options. Adv. Ther. 42, 5879–5895 (2025).

8. Ediae, G. U. et al. ThinkRare: A search algorithm to identify patients with undiagnosed rare genetic disease in an electronic medical record. Genet. Med. 27, 101570 (2025).

9. Lundberg, S. M., Erion, G. G. & Lee, S.-I. Consistent individualized feature attribution for tree ensembles. arXiv [cs.LG] (2018).

